# The efficacy and safety of monoclonal antibody treatments against COVID-19: A systematic review and meta-analysis of randomized clinical trials

**DOI:** 10.1101/2021.06.04.21258343

**Authors:** Ifan Ali Wafa, Nando Reza Pratama, David Setyo Budi, Henry Sutanto, Alfian Nur Rosyid, Citrawati Dyah Kencono Wungu

**Affiliations:** Faculty of Medicine, Universitas Airlangga, Indonesia; Department of Cardiology, CARIM School for Cardiovascular Diseases, Maastricht University, The Netherlands; Department of Physiology and Biophysics, State University of New York (SUNY) Downstate Health Sciences University, New York, USA; Department of Pulmonology and Respiratory Medicine, Faculty of Medicine, Universitas Airlangga, Indonesia; Department of Physiology and Medical Biochemistry, Faculty of Medicine, Universitas Airlangga, Indonesia; Institute of Tropical Disease, Universitas Airlangga, Indonesia

**Keywords:** COVID-19, Monoclonal Antibody, Mortality, Viral load, Meta-analysis

## Abstract

**Objectives:** The use of monoclonal antibody for COVID-19 showed conflicting results in prior studies and its efficacy remains unclear. We aimed to comprehensively determine the efficacy and safety profile of monoclonal antibodies in COVID-19 patients.

**Methods:** Sixteen RCTs were analyzed using RevMan 5.4 to measure the pooled estimates of risk ratios (RRs) and standardized mean differences (SMDs) with 95% CIs.

**Results:** The pooled effect of monoclonal antibodies demonstrated mortality risk reduction (RR=0.89 (95%CI 0.82-0.96), I^2^=13%, fixed-effect). Individually, tocilizumab reduced mortality risk in severe to critical disease (RR=0.90 (95%CI 0.83-0.97), I^2^=12%, fixed-effect)) and lowered mechanical ventilation requirements (RR=0.76 (95%CI 0.62-0.94), I^2^=42%, random-effects). Moreover, it facilitated hospital discharge (RR=1.07 (95%CI 1.00-1.14), I^2^=60%, random-effects). Meanwhile, bamlanivimab-etesevimab and REGN-COV2 decrease viral load ((SMD=-0.33 (95%CI -0.59 to -0.08); (SMD=-3.39 (95%CI -3.82 to -2.97)). Interestingly, monoclonal antibodies did not improve hospital discharge at day 28-30 (RR=1.05 (95%CI 0.99–1.12), I^2^=71%, random-effects) and they displayed similar safety profile with placebo/standard therapy (RR=1.04 (95%CI 0.76-1.43), I^2^=54%, random-effects).

**Conclusion:** Tocilizumab improved hospital discharge and reduced mortality as well as the need for mechanical ventilation, while bamlanivimab-etesevimab and REGN-COV2 reduced viral load in mild to moderate outpatients. In general, monoclonal antibodies are safe and should be considered in severe to critical COVID-19 patients.

**Registration:** PROSPERO (CRD42021235112)

## INTRODUCTION

Since December 2019, a novel coronavirus disease (COVID-19) firstly discovered in Wuhan, China has spread globally and profoundly affected various aspects of life (Li et al., 2020). The viral infectious disease is caused by SARS-CoV-2; an enveloped, positive-sense, single-stranded genomic ribonucleic acid (+ssRNA) virus from the group of *Betacoronavirus* in the family of *Coronaviridae* (Hu et al., 2021). In the lungs, SARS-CoV-2 binds to angiotensin converting enzyme type-2 (ACE-2) receptors at the membrane of pulmonary alveolar cells type-2 and undergoes endocytosis. Subsequently, the interaction of viral antigen with RIG-I-like receptors (RLRs) activates the host immune system as an effort to eliminate the virus from the body (Hertanto et al., 2021), predisposing to the clinical presentations of COVID-19 patients, ranging from asymptomatic or mild up to severe disease state with pneumonia and acute respiratory distress syndrome that can ultimately lead to death (Lai et al., 2020).

The development of optimal and effective therapies for COVID-19 is essential to minimize COVID-19 morbidity and mortality (Lu, 2020; Li and De Clercq, 2020). Several components of the virus and host immune system have been identified as potential targets in COVID-19 management. A previous study reported that the SARS-CoV-2 S2 protein was important for viral entry and thought to be a potential target for neutralizing antibody (Walls et al., 2020). Moreover, the SARS-CoV-2 infection could trigger a hyperactive immune response, leading to cytokine release syndrome (CRS) or cytokine storm (Hertanto et al., 2021). Among numerous proinflammatory cytokines involved in CRS, interleukin (IL)-6 is one of the most critical and has been associated with a poor prognosis (Zhang et al., 2020a; Zhang et al., 2020b; Zhao, 2020). Therefore, the inhibition of IL-6 (e.g., by preventing the binding to its receptors) could prevent the occurrence of CRS and lower the severity of the disease. Moreover, complement C5a and white blood cells (i.e., neutrophil and monocytes) were detected in the bronchoalveolar lavage fluid (BALF) of COVID-19 patients, supporting the chemoattraction role of C5a in lungs-derived C5aR1-expressing cells; which is responsible for cell damage and ARDS (Carvelli et al., 2020). Of note, C5a is one of the major drivers for complement-mediated inflammation that rapidly responds to pathogens and cellular injury (Woodruff and Shukla, 2020).

Monoclonal antibody is one of the proposed therapeutic options for COVID-19. Anti-SARS-CoV-2 monoclonal antibodies are among the latest investigational COVID-19 treatments granted with emergency use authorization (EUA) from the United States Food and Drug Administration (FDA). Briefly, monoclonal antibodies recognize one epitope of an antigen while polyclonal antibodies recognize multiple epitopes (Lipman et al., 2005). The variable region can be modified to target specific molecules, including the S2-protein, cytokines, and cytokine receptors. Among 5 Antibody isotypes—IgA (subclasses IgA1 and IgA2), IgE, IgD, IgM, and IgG (subclasses IgG1, IgG2, IgG3 and IgG4) —IgG is commonly selected for therapeutic purposes due to its strong binding affinity to an antigen and its Fc receptor, supported by its long serum half-life (Chames et al., 2009; Lu, 2020). As the consequence, the administration of neutralizing monoclonal antibody targeting SARS-CoV-2 spike proteins allows the inhibition of virus attachment to human ACE-2 receptors, thus inhibits viral entry (Tian et al., 2020). To prevent complement system activation triggered during SARS-CoV-2 infection, a recent study proposed the use of monoclonal antibody against C5a (anti-C5a) (Woodruff and Shukla, 2020). Among available monoclonal antibodies for COVID-19, anti-IL-6 receptors and anti-SARS-CoV-2 are widely studied in clinical trials (Yang et al., 2020; Patel et al., 2021).

Nonetheless, the efficacy and safety of this pharmacological agent remain controversial (FDA, 2020; Patel et al., 2021). Moreover, at present, the application of monoclonal antibody as a therapeutic agent in COVID-19 shows conflicting results in prior studies, demanding further investigations. Thus, this meta-analysis aims to assess the previously reported efficacy and safety of monoclonal antibodies on clinical and laboratory outcomes and its safety profile in COVID-19 patients.

## MATERIALS AND METHODS

### Search Strategy

The PubMed (MEDLINE), ScienceDirect, Cochrane Library, Proquest and Springer databases were systematically searched from January 25 until February 5, 2021, without any limitation of publication year. We also performed manual searches, extended from February 5 to March 5, 2021, through MedRxiv and citation searching to get evidence from unpublished data and retrieve potential articles without missing any additional eligible studies. The following keywords were used: “(COVID-19) AND ((Monoclonal Antibody) OR (Neutralizing Antibody) OR (Serotherapy)) AND ((Viral Load) OR (Oxygen) OR (Duration) OR (Mortality) OR (Inflammation))”. Additional details about the search strategy are available in ***Supplementary Materials***.

### Data Collection

The title and abstract of the articles were screened by IAW and NRP. Duplications were removed using the Mendeley reference manager. We independently screened the title and abstract of all retrieved studies based on the following eligibility criteria: (1) participants confirmed at any clinical stage of COVID-19 with/without other comorbidities; (2) adult (≥18 years) male/female study population; (3) the study involved monoclonal antibody treatments of interest; (4) the study compared the intervention group with control (placebo or/and standard of care or combination therapy); (5) the study evaluated efficacy (i.e. mortality, need for mechanical ventilation, hospital discharge, virologic outcomes) or safety outcomes (serious adverse events); (6) study type was randomized controlled trial (RCT).

### Data Extraction and Quality Assessment

IAW, NRP, and DSB independently extracted relevant data using the standardized form. The following information was extracted: first author’s name and publication year, study design, country, sample size, age, disease severity, dosage and administration of monoclonal antibodies, types of comparison, and outcomes (all-cause mortality, need for mechanical ventilation, hospital discharge at day 28-30, change of viral load, and serious adverse events). Serious adverse events were defined as any untoward medical occurrence that are potentially related to monoclonal antibody treatment.

The studies were classified into “low risk of bias,” “some concerns,” or “high risk of bias” according to the Cochrane risk of bias tool for randomized trial (RoB ver.2) (Sterne et al., 2019). Any discrepancies were consulted with an expert and resolved by discussion until reaching consensus. The Grading of Recommendation Assessment, Development, and Evaluation (GRADE) system was used to evaluate the quality of evidence of the findings (Brignardello-Petersen et al., 2018; Puhan et al., 2014).

### Statistical Analysis

Primary analyses were carried out using the Review Manager version 5.4 (The Cochrane Collaboration). Pooled risk ratios (RRs) for dichotomous outcomes were evaluated using Mantel-Haenszel method. Standardized mean differences (SMDs) of continuous outcomes were pooled using inverse variance. *I*^*2*^ test was used to quantify heterogeneity between studies, with values *I*^*2*^>50% represents moderate-to-high heterogeneity. If the value of *I*^*2*^ statistics was <50% or the *p*-value was >0.1, the fixed-effects model could be applied; otherwise, the random-effects model would be used. Begg’s funnel plot and Egger’s test were performed for publication bias analysis, and if present, trim-and-fill method was performed. All statistical analysis with a *p*-value<0.05 was considered statistically significant.

Subgroup analyses were done on monoclonal antibody types and disease severity for mortality risk, and monoclonal antibody types for the other outcomes. Leave-one-out sensitivity analysis was conducted to find the source of statistical heterogeneity and demonstrate how each study affected the overall result. Fixed-effects and random-effects with different tau estimators (DL, SJ, and HKSJ) were performed for sensitivity analysis using R version 4.0.5 to find the robustness of pooled data (see ***Supplementary Materials***).

## RESULTS

### Study characteristics

We identified 6032 and 7310 studies through primary database and manual searching, respectively. After duplication removal, we screened potentially relevant studies and obtained 228 studies to be checked for eligibility. Some studies were excluded due to the reasons documented in PRISMA diagram (***Figure 1***).

**Figure 1.**
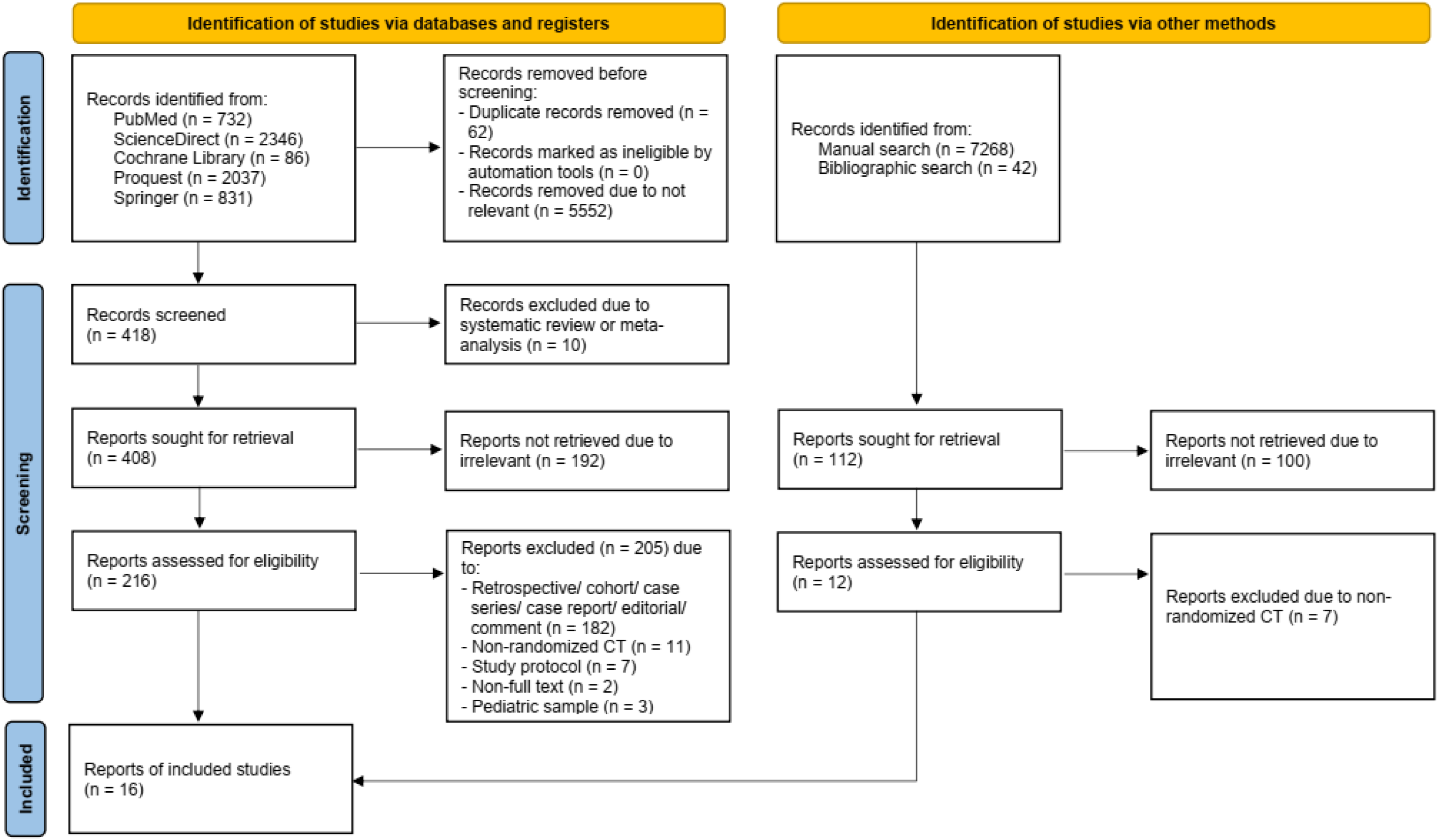
PRISMA diagram of the literature search

Among 16 RCTs, the REMAP-CAP trial (Gordon et al., 2021) was split into two separate intervention groups: the tocilizumab and sarilumab groups. In total, there were 8857 participants included in the meta-analysis, consisted of 4700 and 4157 participants in the intervention and control groups, respectively. The characteristics and outcomes summary for each study, RoB ver.2 assessment, and the certainty of evidence of findings reported using GRADE system are presented in ***Supplementary Tables***.

## MORTALITY

### All-cause mortality

All-cause mortality was examined from 16 RCTs with a total of 8857 patients. Monoclonal antibody was associated with a lower mortality risk (RR=0.89 (95%CI 0.82-0.96), I^2^=3%, fixed-effects). Subsequently, subgroup analyses on the disease severity and monoclonal antibody types were conducted, however only tocilizumab and sarilumab therapies for severe-critical COVID-19 patients were pooled, due to limited studies available (***Figure 2***).

**Figure 2.**
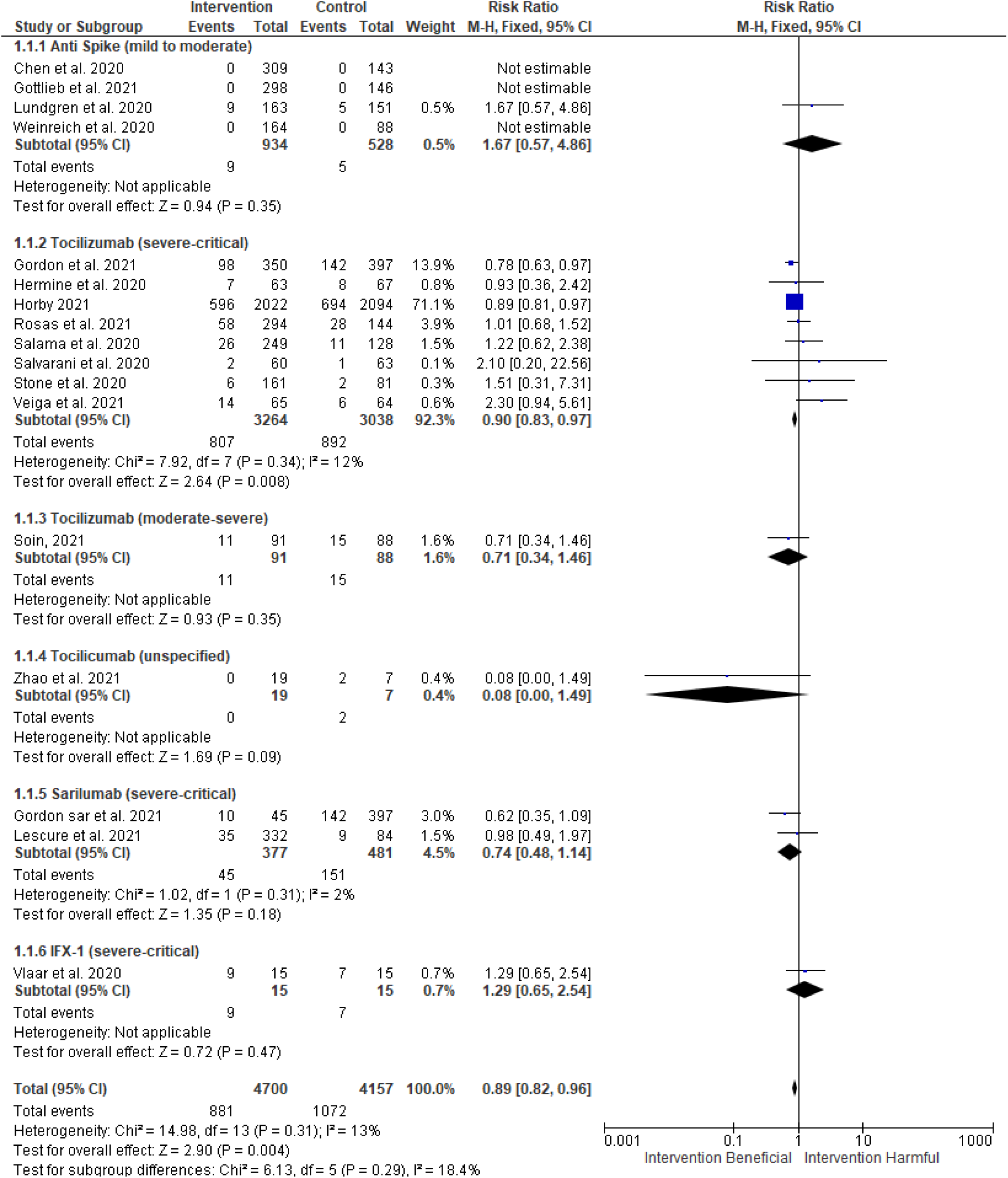
Subgroup analysis between types of monoclonal antibody and mortality among COVID-19 patients

### Tocilizumab in severe-critical COVID-19

Patients with severe-critical COVID-19 receiving tocilizumab displayed a lower mortality risk (RR=0.90 (95%CI 0.83-0.97), I^2^=12%, fixed-effect). RECOVERY trial contributed to the most weight in this meta-analysis (71.1%), and 82.3% in tocilizumab arm and 82.2% in standard therapy arm receiving corticosteroid (pooled RR=0.87 (95%CI 0.80-0.95), I^2^=14%, fixed-effects). Omitting this trial did not change the direction of effect, although it impaired the statistical significance (RR=0.92 (95%CI 0.78-1.10), I^2^=25%, fixed-effects). Meanwhile, TOCIBRAS trial mainly contributed to the statistical heterogeneity. Excluding this study from the analysis provided a more consistent result (pooled RR=0.89 (95%CI 0.82-0.96), I^2^=0%). At last, Funnel plot and Egger’s test did not show any publication bias.

### Sarilumab in severe-critical COVID-19

In severe to critical COVID-19 patients treated with sarilumab, the pooled effect was evaluated using two studies: EudraCT and REMAP-CAP trials (RR=0.74 (95%CI 0.48-1.14), I^2^=2%, fixed-effect). Numerically, the RR was lower than that of tocilizumab, but it did not reach statistical significance. Egger’s test cannot be performed because there were only two studies included in the analysis.

## THE NEED FOR MECHANICAL VENTILATION

Ten RCTs consisted of 6061 patients were examined (***Figure 3***). Only studies involving anti-IL-6R antibodies reported mechanical ventilation outcome. All participants were in severe-critical disease, except for the participants from COVINTOC trial who were in moderate-severe state. Although IL-6R did not demonstrate a reduction of mechanical ventilation requirements (RR=0.76 (95%CI 0.61-0.96, I^2^=49%, random-effects), subgroup analyses revealed that tocilizumab significantly reduced the need for mechanical ventilation (RR=0.76 (95%CI 0.62-0.94), I^2^=42%, random-effects), but not sarilumab (RR=0.76 (95%CI 0.21-2.78), I^2^=87%, random-effects). We also found that EMPACTA trial was the source of heterogeneity in the tocilizumab subgroup. However, omitting this study yielded a similar result (RR=0.81 (95%CI 0.72-0.91), I^2^=0%, fixed-effects). No publication bias was detected from the funnel plot and Egger’s test.

**Figure 3.**
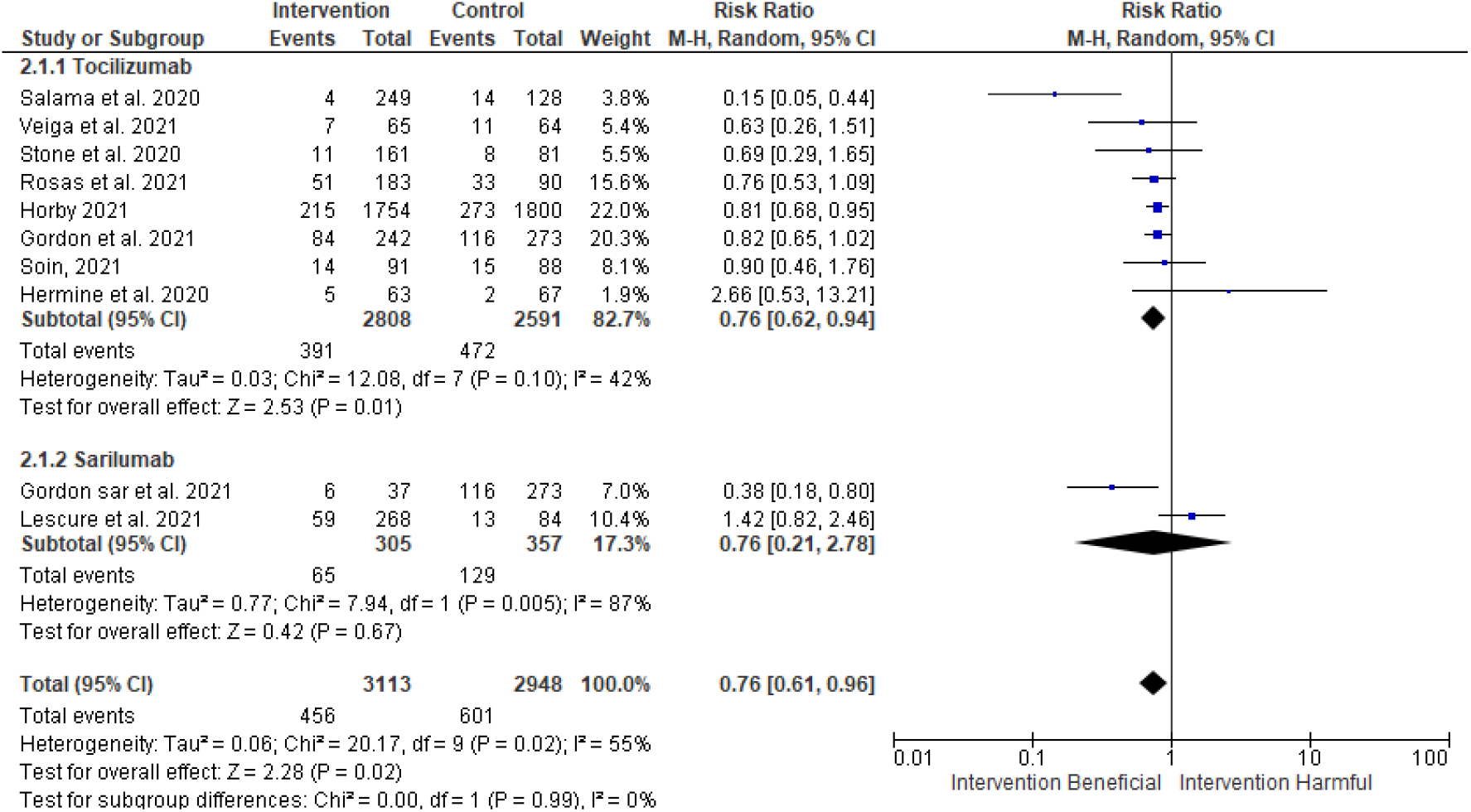
Subgroup analysis between types of monoclonal antibody and the need for mechanical ventilation among COVID-19 patients

## HOSPITAL DISCHARGE AT DAY 28-30

Eleven RCTs consisted of 7490 patients were examined (***Figure 4***). The overall effect of the interventions on hospital discharge at day 28-30 showed no significant difference (RR=1.05 (95%CI 0.99-1.22), I^2^=71%, random-effects). Patients in tocilizumab and sarilumab subgroups had severe-critical disease, while patients receiving spike-protein antibodies were in moderate-severe COVID-19. Subsequently, subgroup analyses for sarilumab and tocilizumab were performed and only tocilizumab significantly increased the rate of hospital discharge (RR=1.07 (95%CI 1.00-1.14), I^2^=60%, random-effects). The *p*-value in 4 significant figures was 0.0498, therefore it reached statistical significance. The funnel plot and Egger’s test did not indicate any publication bias.

**Figure 4.**
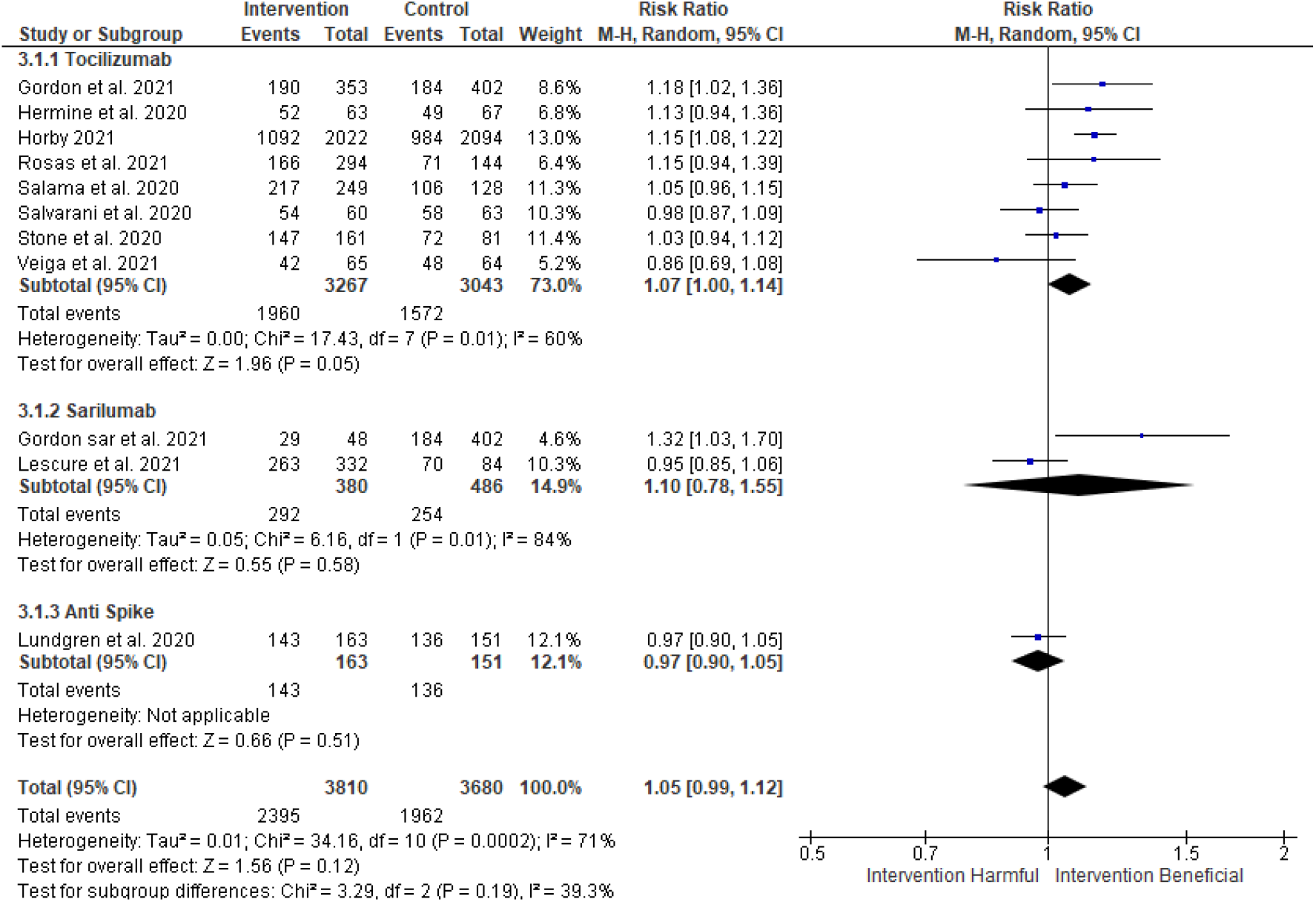
Subgroup analysis between types of monoclonal antibody and the number of COVID-19 patients discharged from hospital at day 28-30

## CHANGE OF VIRAL LOAD

Two RCTs consisted of 896 patients with bamlanivimab monotherapy (***Figure 5***) indicated that bamlanivimab alone did not reduce viral load at day 11 (SMD=-0.07 (95%CI -0.21 to 0.07), I^2^=44%, fixed-effect), in contrast to the combination of bamlanivimab and etesevimab (SMD=-0.33 (95%CI -0.59 to -0.08)). In addition, REGN-CoV2 significantly reduced viral load at day 7 (SMD=-3.39 (95%CI -3.82 to -2.97)). However, only two studies included into the analysis, thus Egger’s test could not be performed.

**Figure 5.**
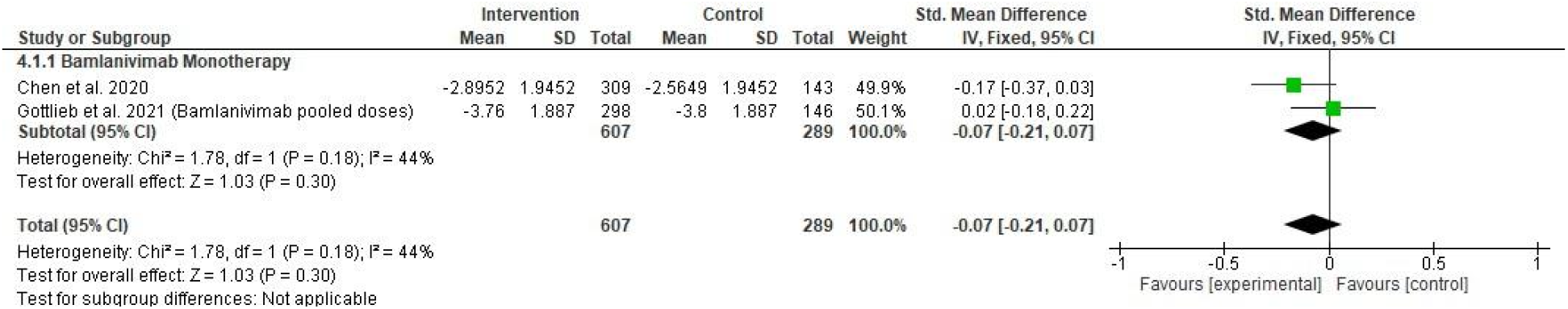
Forest plot for the correlation between monoclonal antibody and viral load change from baseline

## SERIOUS ADVERSE EVENTS

In 16 RCTs of 8897 patients, the overall safety of monoclonal antibody did not differ from placebo/standard therapy (RR=1.04 (95%CI 0.76-1.43), I^2^=54%, random-effects) (***Figure 6***), as well as the subgroup analyses performed for anti-spike-protein (RR=1.00 (95%CI 0.67-1.49) I^2^=29%, fixed-effect), tocilizumab (RR=0.96 (95%CI 0.79-1.18), I^2^=72%, random-effects) and sarilumab (RR=1.12 (95%CI 0.74-1.70, I^2^=0%, fixed-effect). Interestingly, the CORIMUNO-TOCI 1 trial reported that tocilizumab was significantly safer than placebo/standard therapy. On the contrary, the EMPACTA trial showed that tocilizumab was more harmful than placebo/standard therapy. These two studies were the main sources of heterogeneity in our analysis, although removing those studies from the tocilizumab subgroup did not change the direction of effect and statistical significance (RR=1.00 (95%CI 0.80-1.24), I^2^=0%, fixed-effects). Publication bias was indicated in the anti-spike-protein subgroup. The trim-and-fill method altered neither the direction of effect nor the statistical significance (RR=0.92 (95%CI 0.63-1.36), I^2^=27%, fixed-effects).

**Figure 6.**
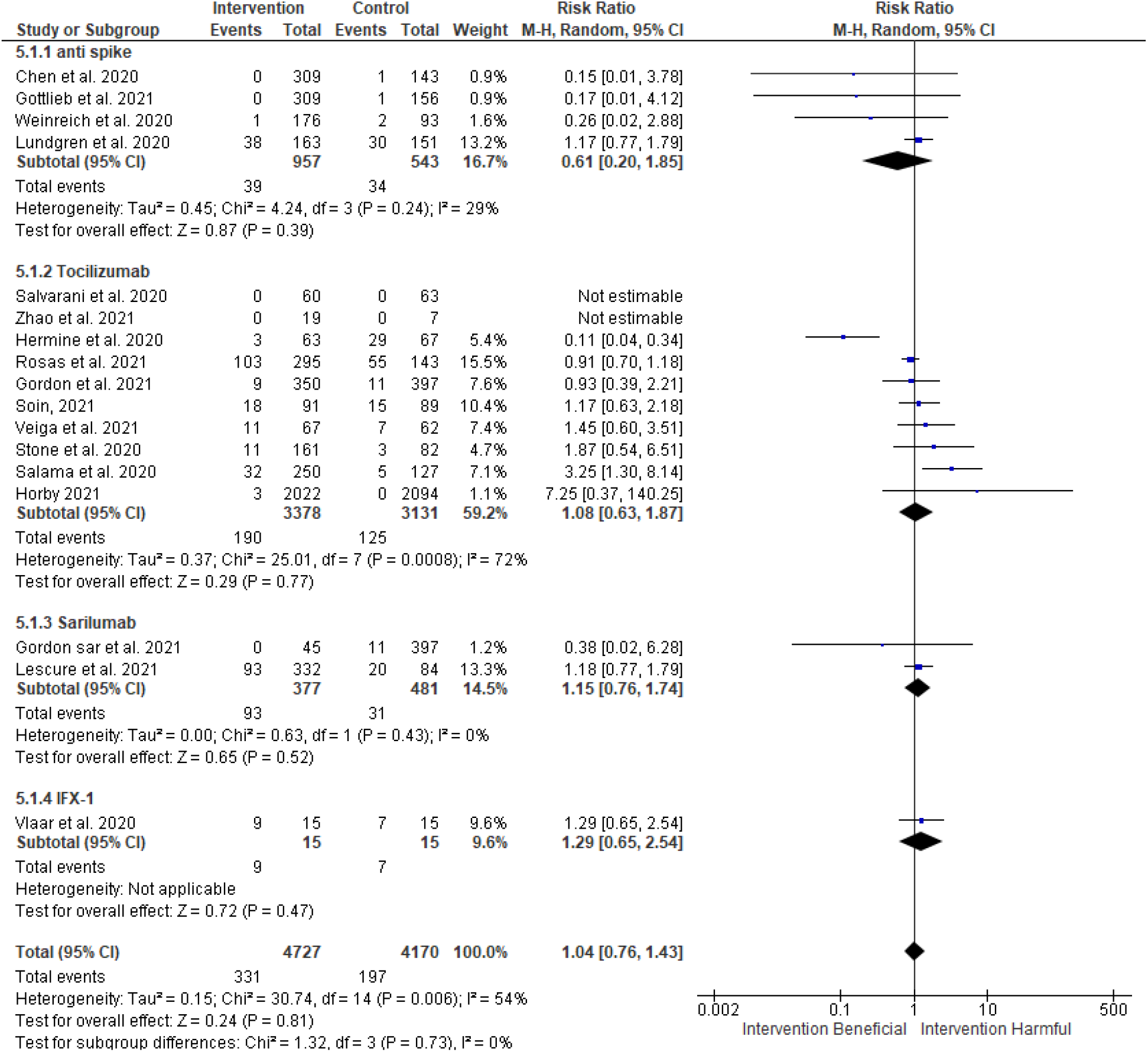
Subgroup analysis between types of monoclonal antibody and the number of serious adverse events in COVID-19

## DISCUSSION

Our analyses showed that monoclonal antibodies provided benefits on mortality rate reduction, mostly because of the weight of tocilizumab studies. From subgroup analysis, tocilizumab showed this benefit, while sarilumab did not. Analysis of the need for mechanical ventilation was conducted by employing only anti-IL-6R studies since the others did not report this outcome. Additionally, some RCTs were outpatients and in mild-moderate disease. No significant benefit was found on hospital discharge at day 28-30 from pooling all monoclonal antibodies. However, subgroup analysis demonstrated that tocilizumab had a significantly higher hospital discharge rate, but sarilumab and bamlanivimab did not. Most of the publications included in this study did not specify any specific description about the hospital discharge. We obtained this data from the description of their ordinal severity scale. In addition, the symptom progression score was not compared because it was described differently across studies.

Next, we also performed a meta-analysis assessing the efficacy of monoclonal antibodies in lowering viral load. For this particular purpose, all included studies employed anti spike-proteins (Chen et al., 2021; Gottlieb et al., 2021; Weinreich et al., 2021). There were three studied interventions—bamlanivimab, bamlanivimab-etesevimab, and REGN-COV2. As the results, the pooled effect of bamlanivimab studies did not show significant viral load reduction. Meanwhile, bamlanivimab-etesevimab and REGN-COV2 reported significant viral load reduction, but each was evaluated only from one RCT, therefore we cannot evaluate the pooled effect. Finally, safety profile should be taken into consideration when administering monoclonal antibodies in COVID-19 patients. Our analyses showed that monoclonal antibodies did not show significant harm or benefit as compared to placebo/standard care. However, not all studies included in this meta-analysis specified the number of treatment-related severe adverse events. The researchers considerately assumed that severe adverse events were related to the treatment unless it was specified otherwise.

We believe that pooling different monoclonal antibodies into one outcome analysis is not insightful since different interventions should be treated independently as they could display varying results. Therefore, we also aimed to look into the subgroup analysis in addition to the overall effect of the pooled antibodies.

## ANTI-IL-6R

The efficacy of tocilizumab was mostly reported in studies where the participants received corticosteroids, for example in RECOVERY and REMAP-CAP trials. If these studies were excluded from the analysis, the pooled effect of mortality risk reduction was no longer significant. This highlights the potential benefit of the combination of tocilizumab and corticosteroids in mortality risk reduction. Indeed, it was shown that dexamethasone significantly reduced mortality risk (Sterne et al, 2020). This could also indicate that the tocilizumab effect may be additional to the corticosteroid benefit (Horby et al, 2021). Whether the administration of tocilizumab alone, without another immunomodulatory agent, would reduce mortality risk remains unclear. Therefore, it is recommended that the use of tocilizumab in severe-critical COVID-19 patients is combined with corticosteroids rather than tocilizumab alone.

Next, our subgroup analysis showed that tocilizumab was beneficial in reducing mechanical ventilation needs and hospital discharge. Consistently, the benefits of tocilizumab in improving oxygen-support and mechanical ventilation rate have been previously documented (Aziz et al., 2021; Kotak et al., 2020; Tleyjeh et al., 2021). Additionally, REMAP-CAP and EMPACTA trials reported that the administration of tocilizumab was associated with earlier hospital discharge and reduction of hospital stay, respectively.

We failed to demonstrate the benefit of sarilumab in reducing the need for mechanical ventilation and hospital discharge rate. This was in line with another study (Della-Torre et al., 2020). The following are some of the proposed arguments to explain this finding. First, the open-label REMAP-CAP trial showed that the sample size included in the sarilumab group was smaller than the control group. Second, the double-blind EudraCT trial reported that more than 60% of patients in the trial received systemic corticosteroids before, during, or after infusion of the studied drug and the frequency of systemic corticosteroid varied during the study. Consequently, it might have diminished the differences between the investigational drug and the control group.

Anti IL-6R was generally safe and tolerable for the treatment of COVID-19 patients. A previous study reported no significant differences between tocilizumab and control groups in terms of the risk of treatment-related serious adverse events (Lin et al, 2021). Tocilizumab, moreover, had a lower rate of serious infections compared to those in the control group (Kotak et al, 2020; Lan et al., 2020; Zhao et al., 2021).

Lastly, the SARS-CoV-2 infection may result in excessive release of pro-inflammatory cytokines, including IL-6, an important cytokine associated with disease severity and mortality, which leads to hyperactivation of the immune response associated with acute respiratory distress syndrome (Xu et al., 2020). However, although IL-6 is one of major cytokines that drive CRS, IL-6 suppression alone might be insufficient to cease the hyperinflammatory phase of COVID-19. Moreover, although there was an increased IL-6 level in COVID-19 patients, it was not as high as observed in sepsis or ARDS (Leisman et al., 2020; Sinha et al., 2020). In general, healthcare providers need to consider patients with severe COVID-19 to attain the maximum benefit from the inhibition of IL-6.

## ANTI-SPIKE-PROTEIN

Viral load reduction of bamlanivimab was evaluated by two studies (Chen et al., 2021; Gottlieb et al., 2021) from the same RCT, BLAZE-1 (NCT04427501). Both studies reported that bamlanivimab 700mg and bamlanivimab 7000mg did not reduce viral load at day 11. Gottlieb et al. (2021) reported that only bamlanivimab 2800mg showed a higher viral load reduction although it was statistically insignificant, while Chen et al. (2020) reported that bamlanivimab 2800mg significantly reduced viral load.

Prozone-like effect possibly plays a role for the reduced efficacy at higher dose, more likely in the earlier time of the disease (Vaidya et al., 2017; Casadevall et al., 2021). Besides, the natural course of the disease also plays a role in viral load reduction. This may mask the clinical significance of neutralizing antibody administration. It is important to mention that missing data from Gottlieb et al. (2020) was replaced using an approach detailed in ***Supplementary Materials***.

Among these anti-spike-protein antibodies, REGN-COV2 and bamlanivimab-etesevimab showed statistically significant viral load reduction but bamlanivimab alone did not reduce viral load. This finding was in line with FDA’s decision published on 16 April 2021 stating that FDA revoked its previously issued EUA for monoclonal antibody bamlanivimab (FDA, 2021). There is a possibility that etesevimab has superior efficacy on viral load reduction. Another possibility is that etesevimab and bamlanivimab may have synergistic effects. However, further research on clinical efficacy of etesevimab monotherapy and its pharmacokinetics with bamlanivimab are warranted.

Finally, it is important to note that BLAZE-1, ACTIV-3 and REGN-COV2 research were sponsored by the same company that designed the drug. Moreover, although all serious adverse events were reported, only some were judged by investigators to be treatment related. However, these studies are double-blinded. To correct the publication bias, trim-and-fill method was performed and the result remained similar.

## ANTI-C5a

At present, there was only one study investigating anti-C5a with two outcomes included in this meta-analysis: the all-cause mortality and safety profile of the monoclonal antibody therapy. The mortality rate in anti-C5a antibody IFX-1 (vilobelimab) group vs. placebo was numerically lower, 13.3% vs 26.7%, respectively. However, this finding did not reach statistical significance, probably owing to the small number of participants. Additionally, compared to placebo/standard care, neither benefit nor harm was observed.

## LIMITATION

This meta-analysis has several limitations: (1) some studies were open-labelled; thus, the risk of bias regarding allocation concealment in those studies could not be ruled out; (2) We mainly discussed IL-6R inhibitors (e.g., tocilizumab) because most published RCTs are tocilizumab-associated studies and existing studies on other antibodies are scarce. (3) The small number of participants in some studies may increase the likelihood of type II statistical error. Larger scale RCTs are required to confirm the findings; (4) many other trials are ongoing; therefore, this study needs to be updated as soon as more trials become available. Nevertheless, this meta-analysis provides a solid evidence by incorporating only RCTs. To the best of our knowledge, this is the first meta-analysis that investigated the efficacy and safety of various monoclonal antibodies on clinical and laboratory outcomes, as well as their safety profile in patients with COVID-19.

## CONCLUSION

In conclusion, monoclonal antibody is beneficial in reducing mortality risk and the need for mechanical ventilation, but not hospital discharge in COVID-19 patients. In contrast to sarilumab, tocilizumab reduces mortality risk in severe to critical patients, reduces the need for mechanical ventilation, and increases hospital discharge at day 28-30. Bamlanivimab monotherapy does not reduce mortality, increase hospital discharge, nor reduce viral load; while bamlanivimab-etesevimab and REGN-COV2 significantly decrease viral load. IFX-1 shows no benefit in mortality risk reduction. No major safety concern was documented for all the monoclonal antibodies.

## Data Availability

Not available

## Data Availability

Not available

## Ethical approval

Not applicable.

