## Supplementary Files for "The efficacy and safety of monoclonal antibody treatments against COVID-19: A systematic review and meta-analysis of randomized clinical trials"

### 1.1. Literature Search

**1.1.1. Keywords**

#1 (COVID-19)

#2 (Monoclonal Antibody) OR (Neutralizing Antibody) OR (Serotherapy)

#3 (Viral Load) OR (Oxygen) OR (Duration) OR (Mortality) OR (Inflammation)

**1.1.2. Search Results**

| Database | Keywords | Search Results | Search-Time |
| --- | --- | --- | --- |
| PubMed | #1 AND #2 AND #3 | 732 | 25/01/2021 to 05/02/2021 |
| Sciencedirect | #1 AND #2 AND #3 | 2346 | 25/01/2021 to 05/02/2021 |
| ProQuest | #1 AND #2 AND #3 | 2037 | 25/01/2021 to 04/02/2021 |
| Springer | #1 AND #2 AND #3 | 831 | 25/01/2021 to 05/02/2021 |
| Cochrane Database | #1 AND #2 AND #3 | 86 | 25/01/2021 to 05/02/2021 |
| Manual Search*  (PubMed, Sciencedirect, ProQuest, Springer, Cochrane Database, MedRxiv, and BioRxiv) | - | 7268 | 05/02/2021 to 05/03/2021 |
| Bibliographic search* | - | 42 | 05/02/2021 to 05/03/2021 |

* We conducted this search according to the name of monoclonal antibodies (restricted to phase 2, phase 3, and phase 4 trials) listed in <https://chineseantibody.org/covid-19-track/>

### 2.1. Calculating missing data

We manually derive the equation for combining variance between two or more population groups. This is meant to get missing pooled-dose data from Gottlieb et al., 2021.

#### 2.1.1. Variance of a population

When set of data in a population is written as the following:

$a_{1}, a_{2}, \ldots, a_{n}$	(1)

Therefore, the mean,$\bar{x}$, is:

$\bar{x}=\frac{a_{1}+...+a_{n}}{n}=\frac{\sum_{i=1}^{n1} a_{i}}{n}$	(2)

Hence, the total sum is:

$\sum_{i=1}^{n1} a_{i}=n \bar{x}$	(3)

The squared deviation for each data is

$\left( a_{i}-\bar{x} \right)^{2}$	(3)

Variance total squared deviation from set of data in an entire population. Therefore, the variance is:

$s^{2}= \frac{{{(a}_{1}-\bar{x})}^{2}+...+{{(a}_{n}-\bar{x})}^{2}}{n}=\frac{\sum_{i=1}^{n1} {{(a}_{i}-\bar{x})}^{2}}{n}$	(4)

#### 2.1.2. Variance of the combined population with algebric approach

If there are N population combined, the set of data can be arranged as the following:

$a_{1},\ldots,a_{n_{1}},b_{1},\ldots,b_{n_{2}},\ldots,n_{1},\ldots,n_{n_{n}}$	(5)

Therefore, the mean,$\bar{x}_{N}$, is:

$\bar{x}_{N}=\frac{a_{1}+\ldots+a_{n1}+b_{1}+\ldots+b_{n2}+n_{1}+\ldots+n_{nn}}{n_{2}+n_{2}+\ldots+n_{n}}=\frac{\sum_{i=1}^{n1} a_{i}+\sum_{i=1}^{b2} b_{i}+...\sum_{i=1}^{nn} n_{n}}{\sum_{i=1}^{n} n}$	(6)

The mean for the entire population data differs from its original subset. Thus, for the entire population, the squared deviation for each data is:

${{(a}_{i}{-\bar{x}}_{N})}^{2}$	(7)

The mean difference between a population of interest and entire population is:

$\bar{x}_{i}{-\bar{x}}_{N}=\Delta m_{i}$	(8)

By a little rearrangement, $\bar{x}_{N}$ can be expressed as:

$\bar{x}_{N}=\bar{x}_{i}-\Delta m_{i}$	(9)

Hence, the squared deviation can be written as the following:

${{(a}_{i}-(\bar{x}_{i}-\Delta m))}^{2}$

$=a_{i}^{2}-2a_{i}\left( \bar{x}_{i}-\Delta m_{i} \right)+(\bar{x}_{i}-\Delta m_{i})^2$

${=a}_{i}^{2}-2a_{i}\bar{x}_{i}+2a_{i}\Delta m_{i}+{\bar{x}_{i}}^{2}-2\bar{x}_{i}\Delta m_{i}+{{\Delta m}_{i}}^{2}$

$=a_{i}^{2}-2a_{i}\bar{x}_{i}+{\bar{x}_{i}}^{2}+2a_{i}\Delta m_{i}-2\bar{x}_{i}\Delta m_{i}+{{\Delta m}_{i}}^{2}$

$=\left( a_{i}-\bar{x}_{i} \right)^{2}+2a_{i}\Delta m_{i}-2\bar{x}_{i}\Delta m_{i}+{{\Delta m}_{i}}^{2}$	(10)

Assume that the mean difference for population a, b, …, n are $\Delta m_{a}$,${\Delta m}_{b}$, …, $\Delta m_{n}$, respectively; and there are $n_{a}$,$n_{b}$, … $n_{n}$data for population a, b, … n, respectively. Therefore, total squared deviation in the entire population is:

$\sum_{i=1}^{n_{a}} \left( a_{i}-\bar{x} \right)^{2}+2a_{i}\Delta m_{a}-2\bar{x}\Delta m_{a}+{{\Delta m}_{a}}^{2})+\ldots+\sum_{i=1}^{n_{n}} \left( n_{i}-\bar{x} \right)^{2}+2a_{i}\Delta m_{n}-2\bar{x}\Delta m_{n}+{{\Delta m}_{n}}^{2}$	(11)

For this Population ‘a’ this part of equation (11) can be expanded as:

$\left( a_{1}-\bar{x} \right)^{2}+\ldots+\left( a_{n_{a}}-\bar{x} \right)^{2}+\Delta m_{a} ((2a_{1}+\ldots+{2 a}_{n_{a}})-2(n_{a}\bar{x})+{(n}_{a}\Delta m_{a}))$	(12)

For simplicity, equation (12) can be written as:

$\sum_{i=1}^{n_{a}} \left( a_{i}-\bar{x} \right)^{2}+\Delta m_{a}\left( \left( \sum_{i=1}^{n_{b}} 2a_{i} \right)-2n_{a}\bar{x}+n_{a}\Delta m_{a} \right)$	(13)

Since $\left( \sum_{i=1}^{n_{b}} 2a_{i} \right)=2n_{a}\bar{x}$ and ${\sum_{i=1}^{n_{b}} \left( a_{i}-\bar{x} \right)}^{2}$= $s_{a}^{2}\times n_{a}$; equation (12) can be simplified as:

$s_{a}^{2}\times n_{a}+\Delta m_{a} ((2n_{a}\bar{x}-2(n_{a}\bar{x})+{(n}_{a}\Delta m_{a}))$

$=s_{a}^{2}\times n_{a}+\Delta m_{a} {(n}_{a}\Delta m_{a})$

${=s}_{a}^{2}\times n_{a}+n_{a}{\Delta m_{a}}^{2}$	(14)

This calculation applies the same for the other population. Hence, the equation (11) can be written as:

$s_{a}^{2}\times n_{a}+n_{a}{\Delta m_{a}}^{2}+s_{b}^{2}\times n_{b}+n_{b}{\Delta m_{b}}^{2}+\ldots+s_{n}^{2}\times n_{n}+n_{n}{\Delta m_{n}}^{2}$

$=n_{a}\left( s_{a}^{2}+{\Delta m_{a}}^{2} \right)+n_{b}\left( s_{b}^{2}+{\Delta m_{b}}^{2} \right)+\ldots+n_{n}\left( s_{n}^{2}+{\Delta m_{n}}^{2} \right)$	(15)

Equation (14) is the total squared deviation. Hence, the variance of combined population is:

$\frac{n_{a}\left( s_{a}^{2}+{\Delta m_{a}}^{2} \right)+n_{b}\left( s_{b}^{2}+{\Delta m_{b}}^{2} \right)+\ldots+n_{n}\left( s_{n}^{2}+{\Delta m_{n}}^{2} \right)}{n_{a}+n_{b}+\ldots+n_{n}}$	(16)

As a result, the standard deviation of combined population is:

${SD}_{combined population}=\sqrt{\frac{n_{a}\left( s_{a}^{2}+{\Delta m_{a}}^{2} \right)+n_{b}\left( s_{b}^{2}+{\Delta m_{b}}^{2} \right)+\ldots+n_{n}\left( s_{n}^{2}+{\Delta m_{n}}^{2} \right)}{n_{a}+n_{b}+\ldots+n_{n}}}$	(17)

#### 2.1.3. Estimating missing standard deviation

Standard deviation can be estimated using 95%CI. We used RevMan 5.4.0 calculator to find the standard deviation between two groups by inputting the 95%CI. However, the 95%CI reported were after comparison. Any data reported by Chen et al., can be probably rounded to the nearest 2 significant figures. Since -2.56 could be anywhere between -2.551 and -2.5649, we filled the input as the following otherwise did not follow the rounding rules:

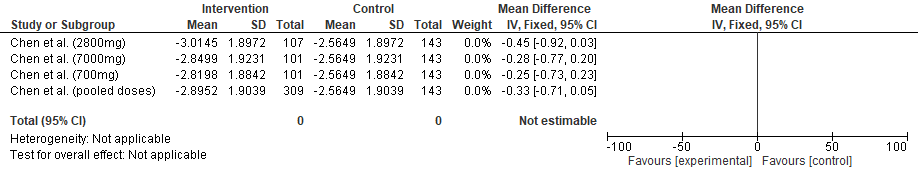

#### 2.1.4. Calculating missing data of pooled doses

Standard deviation for pooled doses is calculated from equation (17):

${SD}_{combined population}=\sqrt{\frac{\left( s_{a}^{2}+{\Delta m_{a}}^{2} \right)+n_{b}\left( s_{b}^{2}+{\Delta m_{b}}^{2} \right)+\ldots+n_{n}\left( s_{n}^{2}+{\Delta m_{n}}^{2} \right)}{n_{a}+n_{b}+\ldots+n_{n}}}$

${SD}_{combined population}=$ 1.9039

This approach has considerable accuracy when tested Chen *et al.* (2020) study. Estimated MD and MD in Chen *et al.* (2020) are -0.33 (95%CI -0.71 to 0.05) and -0.33 (95%CI -0.72 to 0.06), respectively.

The same approach was used for filling the missing standard deviation in Gottlieb et al. (2020) study to estimate RR and 95%CI of the pooled doses.

### 3.1. Sensitivity analysis

| **Outcome** | **Methods** | **Measurement effects** | | **Heterogeneity** | | | **Egger’s test (p)** | **Publication bias** |
| --- | --- | --- | --- | --- | --- | --- | --- | --- |
|  |  | Effect Sizes (95%CI) | p | I^2^ | tau^2^ | P |  |  |
| Mortality (pooled monoclonal antibodies) | DL, fixed | 0.8866 [0.8198; 0.9589] | 0.0026 | 10.2% | 0.0055 | 0.3414 | 0.652626 | No indication |
|  | DL, random | 0.8863 [0.7848; 1.0009] | 0.0518 | 10.2% | 0.0055 | 0.3414 | 0.652626 | No indication |
|  | SJ, random | 0.9518 [0.6763; 1.3396] | 0.7769 | 10.1% | 0.2351 | 0.3419 | 0.652626 | No indication |
|  | HKSJ, random | 0.9518 [0.7192; 1.2596] | 0.7093 | 10.1% | 0.2351 | 0.3419 | 0.652626 | No indication |
| Mortality (tocilizumab; severe-critical) | DL, fixed | 0.8972 [0.8278; 0.9724] | 0.0083 | 11.7% | 0.0043 | 0.3393 | 0.09058772 | No indication |
|  | DL, random | 0.9017 [0.7980; 1.0190] | 0.0972 | 11.7% | 0.0043 | 0.3393 | 0.09058772 | No indication |
|  | SJ, random | 1.0010 [0.7643; 1.3109] | 0.9944 | 11.4% | 0.0630 | 0.3417 | 0.09058772 | No indication |
|  | HKSJ, random | 1.0010 [0.7733; 1.2956] | 0.9932 | 11.4% | 0.0630 | 0.3417 | 0.09058772 | No indication |
| Mortality (sarilumab; severe-critical) | DL, fixed | 0.7417 [0.4806; 1.1446 | 0.1770 | 2.1% | 0.0022 | 0.3122 | N/A | N/A |
|  | DL, random | 0.7466 [0.4801; 1.1610] | 0.1946 | 2.1% | 0.0022 | 0.3122 | N/A | N/A |
|  | SJ, random | 0.7549 [0.4528; 1.2586] | 0.2810 | 2.0% | 0.0357 | 0.3123 | N/A | N/A |
|  | HKSJ, random | 0.7466 [0.0427; 13.0565] | 0.4327 | 2.0% | 0.0357 | 0.3123 | N/A | N/A |
| Mechanical ventilation (pooled monoclonal antibodies) | DL, fixed | 0.7904 [0.7020; 0.8898] | < 0.0001 | 43.2% | 0.0244 | 0.0902 | 0.4984341 | No indication |
|  | DL, random | 0.7171 [0.4416; 1.1645] | 0.0147 | 43.2% | 0.0244 | 0.0902 | 0.4984341 | No indication |
|  | SJ, random | 0.6379 [0.4325; 0.9407] | 0.1789 | 43.1% | 0.3531 | 0.0913 | 0.4984341 | No indication |
|  | HKSJ, random | 0.7171 [0.4221; 1.2181] | 0.1813 | 43.1% | 0.3531 | 0.0913 | 0.4984341 | No indication |
| Hospital discharge at day 28-30 (tocilizumab) | DL, fixed | 1.1211 [1.0735; 1.1708] | < 0.0001 | 59.8% | 0.0049 | 0.0148 | 0.3832322 | No indication |
|  | DL, random | 1.0687 [1.0001; 1.1421] | 0.0498 | 59.8% | 0.0049 | 0.0148 | 0.3832322 | No indication |
|  | SJ, random | 1.0681 [0.9954; 1.1461] | 0.0670 | 50.8% | 0.0059 | 0.0473 | 0.3832322 | No indication |
|  | HKSJ, random | 1.0681 [0.9902; 1.1521] | 0.0788 | 50.8% | 0.0059 | 0.0473 | 0.3832322 | No indication |
| Hospital discharge at day 28-30 (sarilumab) | DL, fixed | 1.0466 [0.9402; 1.1651] | 0.4047 | 83.8% | 0.0510 | 0.0130 | N/A | N/A |
|  | DL, random | 1.1001 [0.7830; 1.5456] | 0.5824 | 83.8% | 0.0510 | 0.0130 | N/A | N/A |
|  | SJ, random | 1.0954 [0.8076; 1.4859] | 0.5580 | 81.7% | 0.0394 | 0.0196 | N/A | N/A |
|  | HKSJ, random | 1.0954 [0.1388; 8.6473] | 0.6748 | 81.7% | 0.0394 | 0.0196 | N/A | N/A |
| Viral load (Bamlanivimab) | DL, fixed | -0.1393 [-0.4076; 0.1290] | 0.3089 | 45.3% | 0.0310 | 0.1765 | N/A | N/A |
|  | DL, random | -0.1420 [-0.5048; 0.2209] | 0.4432 | 45.3% | 0.0310 | 0.1765 | N/A | N/A |
|  | SJ, random | -0.1420 [-0.5093; 0.2253] | 0.4485 | 45.3% | 0.0327 | 0.1765 | N/A | N/A |
|  | HKSJ, random | -0.1420 [-2.4942; 2.2102] | 0.5834 | 45.3% | 0.0327 | 0.1765 | N/A | N/A |
| Serious Adverse Event (pooled monoclonal antibodies) | DL, fixed | 1.1044 [1.0629; 1.1475] | < 0.0001 | 70.7% | 0.0076 | 0.0002 | 0.9276446 | No indication |
|  | DL, random | 1.0532 [0.9866; 1.1244] | 0.1199 | 70.7% | 0.0076 | 0.0002 | 0.9276446 | No indication |
|  | SJ, random | 1.0536 [0.9847; 1.1273] | 0.1301 | 62.5% | 0.0084 | 0.0030 | 0.9276446 | No indication |
|  | HKSJ, random | 1.0536 [0.9824; 1.1300] | 0.1274 | 62.5% | 0.0084 | 0.0030 | 0.9276446 | No indication |
| Serious Adverse Event (anti spike-protein) | DL, fixed | 1.0024 [0.6721; 1.4949] | 0.9908 | 29.2% | 0.4539 | 0.2367 | 0.000573617 | Indicated |
|  | DL, random | 0.6138 [0.2037; 1.8500] | 0.3859 | 29.2% | 0.4539 | 0.2367 | 0.000573617 | Indicated |
|  | SJ, random | 0.6220 [0.2096; 1.8457] | 0.3922 | 28.4% | 0.4342 | 0.2419 | 0.000573617 | Indicated |
|  | HKSJ, random | 0.6220 [0.1282; 3.0172] | 0.4092 | 28.4% | 0.4342 | 0.2419 | 0.000573617 | Indicated |
| Serious Adverse Event (tocilizumab) | DL, fixed | 0.9644 [0.7853; 1.1842] | 0.7291 | 72.0% | 0.3737 | 0.0008 | 0.6220066 | No indication |
|  | DL, random | 1.0838 [0.6281; 1.8701] | 0.7726 | 72.0% | 0.3737 | 0.0008 | 0.6220066 | No indication |
|  | SJ, random | 1.1129 [0.5088; 2.4343] | 0.7889 | 72.0% | 0.1551 | 0.0008 | 0.6220066 | No indication |
|  | HKSJ, random | 1.1291 [0.7195; 1.7719] | 0.7883 | 72.0% | 0.1551 | 0.0008 | 0.6220066 | No indication |
| Serious Adverse Event (sarilumab) | DL, fixed | 1.1209 [0.7396; 1.6988] | 0.5905 | 0.0% | 0 | 0.4315 | N/A | No indication |
|  | DL, random | 1.1479 [0.7580; 1.7385] | 0.5147 | 0.0% | 0 | 0.4315 | N/A | No indication |
|  | SJ, random | 1.0737 [0.4688; 2.4592] | 0.8664 | 0.0% | 0.1487 | 0.4362 | N/A | No indication |
|  | HKSJ, random | 1.0737 [0.0214; 53.9019] | 0.8556 | 0.0% | 0.1487 | 0.4362 | N/A | No indication |

The pooled effects were calculated using Mantel-Haenszel for binary outcomes and Inverse variance for continuous outcome. The result obtained from Revman 5.4.1 used DerSimonian-Laird to estimate tau^2^. Sensitivity analysis was conducted here through R version 4.0.5. We tabulated the result of some different calculation methods for comparison. MH-DL: Mantel-Haenszel with DerSimonian-Laird tau^2^ estimator (Default in Revman 5.4.1); MH-SJ: Mantel-Haenszel with Sidik Jonkman tau^2^ estimator; MH-HKSJ: Mantel-Haenszel and Hartung-Knapp adjustment with Sidik Jonkman tau^2^ estimator.
