## Supplementary Tables for "The efficacy and safety of monoclonal antibody treatments against COVID-19: A systematic review and meta-analysis of randomized clinical trials"

**Table 1A.** Characteristics of the included studies

| Reference | Study design | Country | Sample size | | Age, y  Mean ± SD or Median (IQR) | | Disease severity | Dosage and administration | Comparison | Risk of bias |
| --- | --- | --- | --- | --- | --- | --- | --- | --- | --- | --- |
|  |  |  | Intervention  (n=4700) | Control  (n=4157) | Intervention | Control |  |  |  |  |
| **Anti IL-6R(Tocilizumab)** | | | | | | | | | | |
| *Gordon et al, 2021  (REMAP-CAP) | RCT, open-label | USA, Australia, Belgium, Canada, Croatia, Germany, Hungary, Ireland, Netherlands, New Zealand, Portugal, Romania, Spain, UK | 350 | 402 | 61.5 ± 12.5 | 61.1 ± 12.8 | Hospitalized, critical illness | Tocilizumab 8mg/kg, max 800mg, IV over period of 1 hour, could be repeated 12-24 h later | Standard of care | Low |
| Hermine et al., 2020  (CORIMUNO-TOCI 1) | RCT, open-label | France | 63 | 67 | 64.0  (57.1-74.3) | 63.3  (57.1-72.3) | Moderate, severe, and critical illness | Tocilizumab 8mg/kg + usual care IV on day 1. Additional fixed dose 400mg was recommended on day 3 if O_2_ requirement was not decreased by more than 50% | Usual care | Low |
| **Horby et al., 2021  (RECOVERY) | RCT, open-label | United Kingdom | 2022 | 2094 | 63.3 ± 13.7 | 63.9 ± 13.6 | Hospitalized, severe illness | Tocilizumab dose was determined by body weight as follows: 800 mg if weight >90kg; 600 mg if weight >65 and ≤90 kg; 400 mg if weight >40 and ≤65 kg; and 8mg/kg if weight ≤40 kg. could be repeated 12-24 h later if not improved | Usual care | Low |
| Rosas et al., 2021  (COVACTA) | RCT, double-blind | Canada, Denmark, France, Germany, Italy, Netherlands, Spain, UK, USA | 294 | 144 | 60.9 ± 14.6 | 60.6 ± 13.7 | Hospitalized, severe illness | Single dose of Tocilizumab 8mg/kg (max 800mg) IV | Placebo + standard of care | Some concerns |
| Salama et al., 2020  (EMPACTA) | RCT, double-blind | USA | 249 | 128 | 56.0 ± 14.3 | 55.6 ± 14.9 | Hospitalized, severe illness, not receiving mechanical ventilation | One or two doses of Tocilizumab 8mg/kg (max 800mg) IV, plus standard of care | Placebo + standard of care | Low |
| Salvarani et al, 2020  (RCT-TCZ) | RCT, open-label | Italy | 60 | 63 | 61.5  (51.5-73.5) | 60.0  (54.0-69.0) | Severe to critical illness | Tocilizumab 8mg/kg (max 800mg) IV within 8 h from randomization followed by second dose after 12 h | Standard of care | Some concerns |
| Soin et al., 2021  (COVINTOC) | RCT, open-label | India | 91 | 88 | 56 (47-63) | 54 (43-63) | Hospitalized, moderate to severe illness | Single dose of Tocilizumab 6mg/kg (max 480mg) IV + standard of care, additional dose could be administered if clinical symptoms worsened or did not show improvement within 12h to 7 days | Standard of care | Low |
| Stone et al., 2020 (BACC Bay) | RCT, double-blind | USA | 161 | 81 | 61.6  (46.4-69.7) | 56.5  (44.7-67.8) | Severe illness | Single dose of Tocilizumab 8mg/kg (max 800mg) IV + standard of care | Placebo + standard of care | Low |
| Veiga et al., 2021  (TOCIBRAS) | RCT, open-label | Brazil | 65 | 64 | 57.4 ± 15.7 | 57.5 ± 13.5 | Severe to critical illness | Single dose of Tocilizumab 8mg/kg (max 800mg) IV + standard of care | Standard of care | Some concerns |
| Zhao et al., 2021 | RCT, open-label | China | 19 | 7 | TCZ only  71 (48-77) | 75 (34-81) | Hospitalized, all clinical spectrum | Tocilizumab 4-8mg/kg (recommended 400mg) added to 100mL 0.9% NaCl, IV more than 1 h, could be repeated if the fever still exists within 24 h | Favipiravir (1600mg b.i.d on 1^st^ day followed by 600mg b.i.d. from 2^nd^ to 7^th^ day | High |
|  |  |  |  |  | TCZ+Favipiravir  75 (34-81) |  |  |  |  |  |
| **Anti IL-6R (Sarilumab)** | | | | | | | | | | |
| *Gordon et al., 2021  (REMAP-CAP) | RCT, open-label | International | 45 | 402 | 63.4 ± 13.4 | 61.1 ± 12.8 | Hospitalized, critical illness | Single dose of Sarilumab  400mg IV | Standard of care | Low |
| Lescure et al., 2021 (EudraCT) | RCT, double-blind | Argentina, Brazil, Canada, Chile, France, Germany, Israel, Italy, Japan, Russia, and Spain | 332 | 84 | Sarilumab 200mg 58.0  (51.0-67.0) | 60.0  (53.0-69.5) | Hospitalized, severe to critical illness | Divided into two groups, Sarilumab 400mg+NaCl 0.9% and Sarilumab 200mg+NaCl 0.9%, IV, second dose could be repeated within 24-48 h after first dose | Placebo  (NaCl 0.9%) | Low |
|  |  |  |  |  | Sarilumab 400mg 58.0  (48.0-67.0) |  |  |  |  |  |
| **Anti-SARS-CoV-2 Spike Protein (Bamlanivimab)** | | | | | | | | | | |
| Chen et al., 2020 (BLAZE-1) | RCT, double-blind | USA | 309 | 143 | 45 (18-86) | 46 (18-77) | Mild to moderate illness | Divided into three groups,  Bamlanivimab 700mg, 2800mg, and 7000mg, single dose IV over 1 h | Placebo | Low |
| Gottlieb et al., 2021 (BLAZE-1) | RCT, double-blind | USA | 298 | 146 | Bamlanivimab  700mg  39 (31-58) | 46 (35-37) | Mild to moderate illness | Divided into three groups, Bamlanivimab 700mg, 2800mg, and 7000mg, single dose IV over 1 h | Placebo | Low |
|  |  |  |  |  | Bamlanivimab  2800mg  45 (31-56) |  |  |  |  |  |
|  |  |  |  |  | Bamlanivimab  7000mg  46 (34-55) |  |  |  |  |  |
| Lundgren et al., 2020 (ACTIV-3) | RCT, double-blind | USA, Denmark, Singapore | 163 | 151 | 63 (50-72) | 59 (48-71) | Hospitalized, mild to moderate illness | Single dose of Bamlanivimab 7000mg IV over 1 h | Placebo | Low |
| **Anti-SARS-CoV-2 Spike Protein (REGN-COV2)** | | | | | | | | | | |
| Weinreich et al., 2020 | RCT, double-blind | USA | 164 | 88 | 43.0  (35.0-52.0) | 45.0  (34.0-54.0) | Non hospitalized, mild illness | Divided into two groups,  Low dose (2.4g) and high dose (8.0g) of REGN-COV2, administered  IV in a 250ml NaCl 0.9% over 1 h | Placebo  (NaCl 0.9%) | Low |
| **Anti C5a (Vilobelimab)** | | | | | | | | | | |
| Vlaar et al., 2020 (PANAMO) | RCT, open-label | Netherlands | 15 | 15 | 58.0 ± 9.0 | 63.0 ± 8.0 | Hospitalized, severe illness | Five doses of Vilobelimab (IFX-1) 800mg IV plus best supportive care were administered on days 1,2,4,8, and 15. 6^th^ dose could be administered at day 22 to patient who were still intubated while additional dose could be given between days 11 dan 13 if any signs of clinical worsening were detected | Best supportive care | Some concerns |

RCT, randomized controlled trial; SD, standard deviation; IV, intravenous; *study analyzed into two separated intervention groups, Tocilizumab and Sarilumab group; **non-peer-reviewed preprint study

**Table 1B.** Outcomes of the individual studies

| Reference | All-cause mortality  N (%) | | Need for mechanical ventilation N (%) | | Hospital discharge at day 28-30 N (%) | | Change of viral load at day 7 (Mean ± SD) | | | Serious adverse events  N (%) | |
| --- | --- | --- | --- | --- | --- | --- | --- | --- | --- | --- | --- |
|  | Intervention | Control | Intervention | Control | Intervention | Control | Intervention | | Control | Intervention | Control |
| **Anti IL-6R (Tocilizumab)** | | | | | | | | | | | |
| *Gordon et al., 2021 (REMAP-CAP) | 98 (28.0) | 142 (35.8) | 84 (34.7) | 116 (42.5) | 190 (53.8) | 184 (45.8) | NR | | NR | 9 (2.6) | 11 (2.8) |
| Hermine et al., 2020 (CORIMUNO-TOCI 1) | 7 (11.1) | 8 (11.9) | 5 (7.9) | 14 (21.9) | 52 (82.5) | 49 (73.1) | NR | | NR | 20 (31.8) | 29 (43.3) |
| **Horby et al., 2021 (RECOVERY) | 596 (29.5) | 694 (33.1) | 215 (12.3) | 273 (15.2) | 1093 (54.05) | 990 (47.3) | NR | | NR | 3 (0.2) | 0 (0.0) |
| Rosas et al., 2021 (COVACTA) | 58 (19.7) | 28 (19.4) | 51 (27.9) | 33 (36.7) | 166 (56.5) | 71 (49.3) | NR | | NR | 103 (34.9) | 55 (38.5) |
| Salama et al., 2020 (EMPACTA) | 26 (10.4) | 11 (8.6) | 4 (1.6) | 14 (10.9) | 217 (87.1) | 106 (82.8) | NR | | NR | 32 (12.8) | 5 (3.9) |
| Salvarani et al., 2020 (RCT-TCZ) | 2 (3.3) | 1 (1.6) | NR | NR | 54 (90.0) | 58 (92.1) | NR | | NR | 0 (0.0) | 0 (0.0) |
| Soin et al., 2021 (COVINTOC) | 11 (12.1) | 15 (17.0) | 14 (15.4) | 13 (14.8) | NR | NR | NR | | NR | 18 (19.8) | 15 (16.9) |
| Stone et al., 2020 (BACC Bay) | 6 (3.7) | 2 (2.5) | 11 (6.8) | 8 (9.9) | 147 (91.3) | 72 (88.9) | NR | | NR | 11 (6.8) | 3 (3.7) |
| Veiga et al., 2021 (TOCIBRAS) | 14 (21.5) | 6 (9.4) | 7 (10.8) | 11 (17.2) | 42 (64.6) | 48 (75.0) | NR | | NR | 11 (16.4) | 7 (11.3) |
| Zhao et al., 2021 | 0 (0) | 2 (28.6) | NR | NR | NR | NR | NR | | NR | 0 (0.0) | 0 (0.0) |
| **Anti IL-6R (Sarilumab)** | | | | | | | | | | | |
| *Gordon et al., 2021 (REMAP-CAP) | 10 (22.2) | 142 (35.8) | 6 (16.2) | 116 (42.5) | 29 (60.4) | 184 (45.8) | NR | | NR | 0 (0.0) | 11 (2.8) |
| Lescure et al., 2021 (EudraCT) | 35 (10.5) | 9 (10.7) | 59 (22.0) | 13 (19.1) | 263 (79.2) | 70 (83.3) | NR | | NR | 93 (28.0) | 20 (23.8) |
| **Anti-SARS-CoV-2 Spike Protein (Bamlanivimab)** | | | | | | | | | | | |
| Chen et al., 2020  (BLAZE-1) | 0 (0.0) | 0 (0.0) | NR | NR | NR | NR | -2.90 ± 1.94 | | -2.56 ± 1.94 | 0 (0.0) | 1 (0.7) |
| Gottlieb et al., 2021  (BLAZE-1) | 0 (0.0) | 0 (0.0) | NR | NR | NR | NR | 700mg group  -3.72 ± 1.70 | Pooled  -3.76 ± 1.89 | -3.80 ± 1.89 | 0 (0.0) | 1 (0.6) |
|  |  |  |  |  |  |  | 2800mg group  -4.08 ± 1.72 |  |  |  |  |
|  |  |  |  |  |  |  | 7000mg group  -3.49 ± 1.72 |  |  |  |  |
|  |  |  |  |  |  |  | Combination group  -4.37 ± 1.70 | |  |  |  |
| Lundgren et al., 2020 (ACTIV-3) | 9 (5.5) | 5 (3.3) | NR | NR | 143 (87.7) | 136 (90.1) | NR | | NR | 38 (23.3) | 30 (19.9) |
| **Anti-SARS-CoV-2 Spike Protein (REGN-COV2)** | | | | | | | | | | | |
| Weinreich et al., 2020 | 0 (0.0) | 0 (0.0) | NR | NR | NR | NR | -1.74 ± 0.11 | | -1.34 ± 0.13 | 1 (0.6) | 2 (2.2) |
| **Anti C5a (Vilobelimab)** | | | | | | | | | | | |
| Vlaar et al., 2020 (PANAMO) | 2 (13.3) | 4 (26.7) | NR | NR | NR | NR | NR | | NR | 9 (60.0) | 7 (46.7) |

NR, not reported; SD, standard deviation; *study analyzed into two separated intervention groups, Tocilizumab and Sarilumab group; **non-peer-reviewed preprint study

**Table 2.** Risk of Bias Assessment (RoB ver.2)

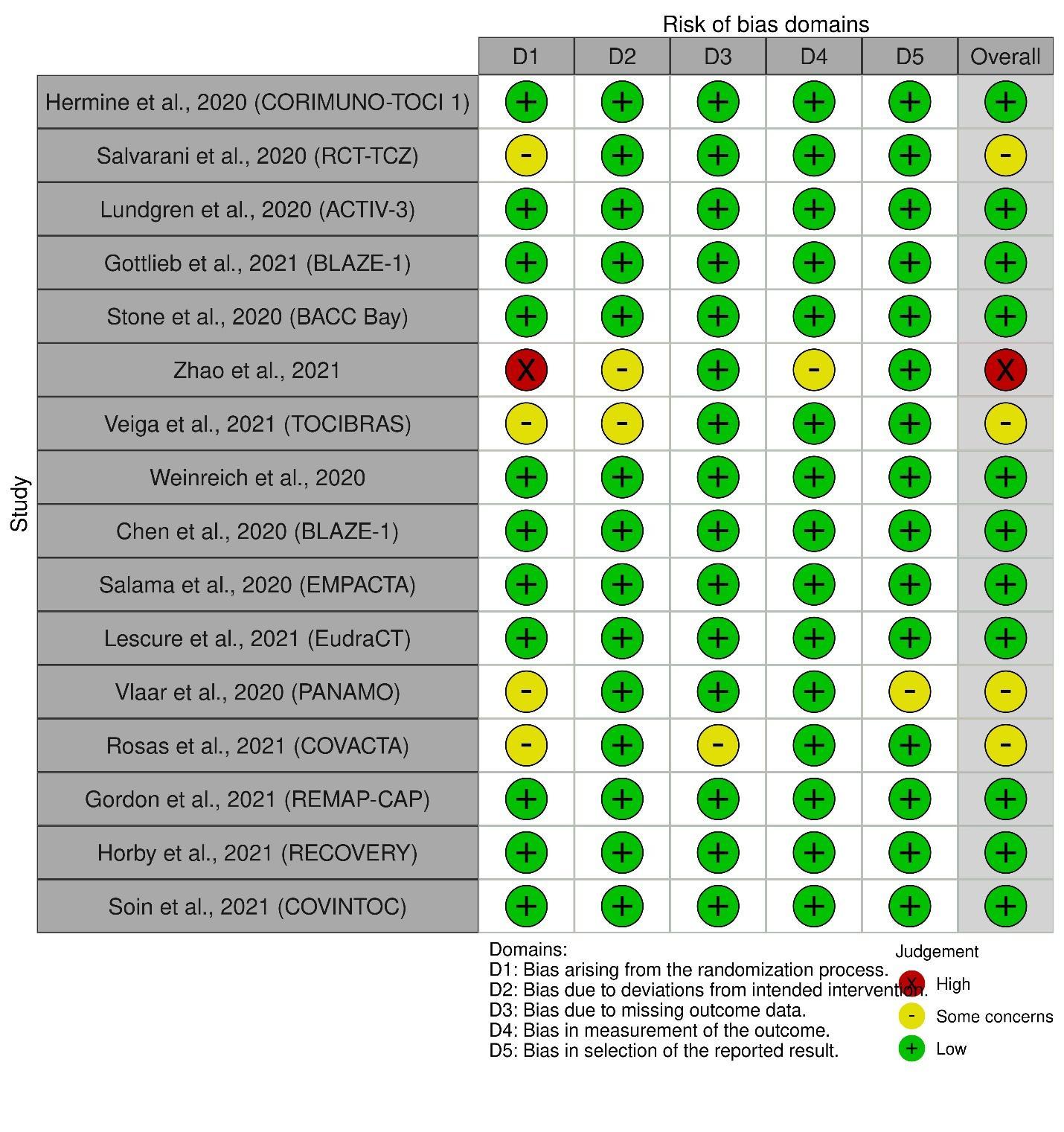

**Table 3.** GRADE Assessment of Evidence

| **Certainty assessment** | | | | | | | **No of patients** | | **Effect** | | **Certainty** | **Importance** |
| --- | --- | --- | --- | --- | --- | --- | --- | --- | --- | --- | --- | --- |
| No of studies | Study design | Risk of bias | Inconsistency | Indirectness | Imprecision | Other considerations | Monoclonal antibody | Standard Care/ Placebo | Relative (95% CI) | Absolute (95% CI) |  |  |
| All-cause Mortality (Pooled: Tocilizumab, Sarilumab, Anti Spike-proteins, IFX-1) | | | | | | | | | | | | |
| 16 | RCT | serious^a^ | not serious | serious^b^ | not serious | none | 881/4700 (18.7%) | 1072/4157 (25.8%) | RR 0.89 (0.82 to 0.96) | **28 fewer per 1,000** (from 46 fewer to 10 fewer) | ⨁⨁◯◯ LOW | IMPORTANT |
| Mortality Risk (Tocilizumab for severe-critical) | | | | | | | | | | | | |
| 8 | RCT | serious^c^ | not serious | serious | not serious | none | 807/3264 (24.7%) | 892/3038 (29.4%) | RR 0.90 (0.83 to 0.97) | **29 fewer per 1,000** (from 50 fewer to 9 fewer) | ⨁⨁◯◯ LOW | CRITICAL |
| Mechanical Ventilation Needs (Pooled: Tocilizumab, Sarilumab) | | | | | | | | | | | | |
| 10 | RCT | serious^d^ | not serious | serious^b^ | not serious | none | 456/3113 (14.6%) | 601/2948 (20.4%) | RR 0.76 (0.61 to 0.96) | **49 fewer per 1,000** (from 80 fewer to 8 fewer) | ⨁⨁◯◯ LOW | CRITICAL |
| Mechanical Ventilation Needs (Tocilizumab) | | | | | | | | | | | | |
| 8 | RCT | serious^c^ | not serious | not serious | not serious | none | 391/2808 (13.9%) | 472/2591 (18.2%) | RR 0.76 (0.62 to 0.94) | **44 fewer per 1,000** (from 69 fewer to 11 fewer) | ⨁⨁⨁◯ MODERATE | CRITICAL |
| Hospital Discharge (Pooled: Tocilizumab, Sarilumab, Bamlanivimab) | | | | | | | | | | | | |
| 11 | RCT | serious^c^ | serious^e^ | serious^b^ | not serious | none | 2538/3810 (66.6%) | 2118/3680 (57.6%) | RR 1.05 (0.99 to 1.11) | **29 more per 1,000** (from 6 fewer to 63 more) | ⨁◯◯◯ VERY LOW | IMPORTANT |
| Viral Load Reduction (Bamlanivimab Monotherapy: Mild-Moderate) | | | | | | | | | | | | |
| 2 | RCT | not serious | serious^e^ | not serious | not serious | none | 607 | 289 | - | SMD 0.07 SD lower (0.24 lower to 0.11 higher) | ⨁⨁⨁◯ MODERATE | CRITICAL |
| Serious Adverse Events (Pooled: Tocilizumab, Sarilumab, Anti Spike-Proteins, IFX-1) | | | | | | | | | | | | |
| 16 | RCT | serious^a^ | not serious^e^ | serious^b^ | serious^f^ | none | 331/4727 (7.0%) | 197/4170 (4.7%) | RR 1.04 (0.76 to 1.43) | **2 more per 1,000** (from 11 fewer to 20 more) | ⨁◯◯◯ VERY LOW | IMPORTANT |

**CI:** Confidence interval; **RR:** Risk ratio; **SMD:** Standardised mean difference; **MD:** Mean difference

**Explanations**

a. Nine RCTs are Open Label; b. Different in intervention; c. Five RCTs are open-label; d. Six RCTs are open-label; e. I^2^ > 30% and P<0.05; f. 95%CI Include null effect (RR = 1) and appreciable harm (RR = 1.25)
